# Validation of diagnosis of acute myocardial infarction and stroke in electronic medical records: a primary care cross-sectional study in Madrid, Spain (the e-MADVEVA Study)

**DOI:** 10.1101/2022.10.01.22280592

**Authors:** C de Burgos-Lunar, I del Cura-González, JC Cárdenas-Valladolid, P Gómez-Campelo, JC Abánades-Herranz, A López-de Andrés, M Sotos-Prieto, V Iriarte-Campo, MA Salinero-Fort

## Abstract

**Objectives:** To validate the diagnoses of AMI and stroke recorded in EMR and to estimate the population prevalence of both diseases in people aged ≥ 18 years

**Design:** Cross-sectional validation study Setting: 45 primary care centres

**Participants:** Simple random sampling of diagnoses of AMI and stroke (ICPC-2 codes K75 and K90, respectively) registered by 55 physicians and random age- and sex-matched sampling of the records that included in primary care EMRs in Madrid (Spain).

**Primary and Secondary Outcomes Measures:** Sensitivity, specificity, positive and negative predictive values, and overall agreement were calculated using the kappa statistic. Applied gold standards were electrocardiograms, brain imaging studies, hospital discharge reports, cardiology reports, and neurology reports. In the case of AMI, the ESC/ACCF/AHA/WHF Expert Consensus Document was also used. Secondary outcomes were the estimated prevalence of both diseases considering the sensitivity and specificity obtained (true prevalence).

**Results:** The sensitivity of a diagnosis of AMI was 98.11% (95% CI, 96.29-99.03), and the specificity was 97.42% (95% CI, 95.44-98.55). The sensitivity of a diagnosis of stroke was 97.56% (95% CI, 95.56-98.68), and the specificity was 94.51% (95% CI, 91.96-96.28). No differences in the results were found after stratification by age and sex (both diseases). The prevalence of AMI and stroke was 1.38% and 1.27%, respectively.

**Conclusion:** The validation results show that diagnoses of AMI and stroke in primary care EMRs constitute a helpful tool in epidemiological studies. The prevalence of AMI and stroke was lower than 2% in the population aged over 18 years.

**Strengths and limitations of this study:**

- The major strength of the e-MADVEVA study is the individual validation (manual validation) of electronic medical records by comparing each case and non-case, matched by age, with an accepted reference gold standard.
- The validation method allows for calculating PPV, NPV, Sensitivity, and Specificity, in contrast with other methods such as questionnaires for healthcare practitioners or patients and comparing rates in a comparable population.
- In-hospital mortality due to AMI and stroke may not have been recorded in primary care EMRs, with the result that the prevalence of patients who die in hospital due to a first AMI or stroke may be underestimated.
- The ICPC-2 codes studied for AMI (ICPC-2 K75) and stroke (ICPC-2 K90) do not allow differentiation between the different types of AMI and stroke.

## Introduction

Electronic medical records (EMR) are digital versions of paper charts in primary care settings and hospitals. EMRs contain notes and information collected by and for clinicians and are used mostly by care providers for diagnosis and treatment. EMRs are more valuable than paper records because they enable providers to track data over time, identify patients for preventive care and screening, monitor patients, and improve the quality of health care (1). In the last 20 years, EMRs have become an increasingly common source of information for research. They allow for more efficient analyses and generalization of findings and minimize selection and recall bias.

Despite these advantages, the validity of some diagnoses recorded in EMRs is uncertain, as the records were not created specifically for research purposes. Errors and inconsistencies in diagnoses can lead to misclassification bias, affecting the quality of research. The EMR must be able to distinguish between those who have had a disease (according to an accepted ‘gold standard’ reference diagnosis) and those who have not.

Health care in Spain is publicly funded and universal. Primary care is usually the gateway to the health system and the point to which most patients treated at other levels of care return (2). In the Community of Madrid, all primary care centres have had EMRs for more than 20 years, and hospital care reports can be accessed. These characteristics make EMRs a valuable source of epidemiological information.

Few studies have validated acute myocardial infarction (AMI) and stroke in primary care EMRs (3) (4) (5) (6) (7) (8). Therefore, our group estimated the validity of the codes for arterial hypertension and diabetes mellitus in primary care EMRs (9) and, more recently, those for atrial fibrillation (10). We obtained high sensitivities, specificities, predictive values, and diagnostic concordance in all of them.

Validation studies determine the degree of systematic measurement error and aid in the interpretation of findings. Researchers increase the reliability of their findings by quantifying the correctness of the data in EMRs.

We chose the diagnoses AMI and stroke because they are major causes of morbidity and mortality and have internationally accepted diagnostic criteria.

This MADrid electronic medical records for Vascular Events VAlidation (e-MADVEVA Study) aims to validate the diagnoses of AMI and stroke recorded in EMR and to estimate the population prevalence of both diseases in people aged ≥ 18 years in the Community of Madrid (Spain).

## Methods

### Design

Cross-sectional validation study of diagnoses of AMI and stroke recorded in primary care EMRs.

### Setting

The e-MADVEVA Study was carried out in 45 primary care centres in the Community of Madrid. These centres provide care to a population of 1,080,000 people. Fifty-five volunteer general practitioners (GPs) took part in the study.

### Sources of information

The e-MADVEVA Study was based on individualized patient data obtained from patients’ primary care EMRs. The EMR, which is managed by the AP-Madrid computer application, is structured around a list of episodes consisting of a code and a description or label. The code corresponds to the second edition of the International Classification of Primary Care (ICPC-2) and can have several different descriptions (11) (12). An episode of care is defined as a health problem or disease from its first presentation to a health care provider to the completion of the last encounter for that same health problem or disease. After several visits by the patient for the same health problem, it is sometimes necessary to change the diagnosis. Therefore, the diagnostic code should be replaced by the definitive one. Unfortunately, the AP-Madrid application allows the ICPC-2 label to be modified without changing the code, thus generating a diagnostic classification error.

### Study population

The e-MADVEVA Study population comprised persons aged ≥ 18 years with active primary care EMRs and at least one entry in the EMR before 1 January 2015.

Patients were not included if they were not regular users, if they were temporarily displaced from their usual residence, or if they were transients who required occasional urgent medical care at the primary care centres.

### Sampling

The sampling procedure has been described elsewhere (9). Briefly, four samples were obtained, two to validate the diagnosis of AMI and two to validate the diagnosis of stroke. Samples 1 and 3 were obtained from clinical records with codes ICPC-2 K-75 (myocardial infarction) and K-90 (stroke), respectively. Samples 2 and 4 were control samples paired by age and sex, without codes K-75 and K-90, respectively.

The participants of samples with codes K-75 and K-90 were selected by random sampling in multistage conglomerates, where the first-stage units were constituted by the GPs and the second-stage units by the patients. The participants of samples without these codes were obtained by individual matching for sex and age at a ratio of 1:1.

Given the absence of reference data on the expected proportion of misclassifications (false negatives or false positives), maximum possible indeterminacy (p=q=0.5) was assumed.

With this assumption, and for a confidence level of 95% and a precision of 5%, 384 patients needed to be assessed for each variable. This was increased to 423 in anticipation of a 10% loss from sampling to validation of the diagnoses (EMR of patients who had become inactive owing to a change of residence, death, or other reasons).

Four patient samples were obtained as follows:

- To validate AMI episodes:

Sample 1: 423 patients with an AMI code (ICPC-2 K75) Sample 2: 423 patients without AMI codes

- To validate stroke episodes:

Sample 3: 423 patients with a stroke code (ICPC-2 K90)

Sample 4: 423 patients without stroke codes.

Given that the probability of presenting these episodes increases with age and that they are is not equally prevalent in both sexes, it was decided that samples 2 and 4 (patients without episodes) should be similar to the respective samples 1 and 3 (patients with episodes) in the variables year of birth and sex. This strategy avoids overestimating specificity if the sample comprised younger patients or was characterized by the predominance of one sex over the other.

Samples 1 and 3 were obtained by simple random sampling from the patient list of participating GPs. Samples 2 and 4 were obtained by individual age and sex matching techniques with their corresponding samples 1 and 3.

Our group collaborates with 153 GPs. Of these, 55 volunteers were selected by random sampling.

A diagnostic test is validated by comparing its results, both positive and negative, with those obtained by the best available instrument for measuring the phenomenon under study (gold standard). In this study, the diagnoses of AMI and stroke in primary care EMRs could be considered equivalent to diagnostic tests.

Patients in samples 1 and 2 were considered to have AMI if they met any of the following criteria:

- Clinical record of hospital discharge report or cardiology outpatient report with a diagnosis of AMI.
- Meeting the diagnostic criteria set out in the third universal definition of AMI established in the ESC/ACCF/AHA/WHF Expert Consensus Document.

Patients in samples 3 and 4 were considered to have had a stroke if they met any of the following criteria:

- Hospital or neurological outpatient discharge report with a diagnosis of stroke
- Sudden onset of a focal neurological deficit, with clinical or imaging evidence of infarction lasting 24 hours or more and not attributable to a nonischemic cause (i.e., not associated with brain infection, trauma, tumor, seizure, metabolic disease, or degenerative neurological disease)
- Acute extravasation of blood into the brain parenchyma or subarachnoid space associated with neurological symptoms.

The evaluators validated the diagnoses by accessing the primary care EMR and, based on the information collected there, verifying that the criteria were met.

The Flow chart of EMR and patients’ selection is shown in Figure 1. The evaluators were GPs with experience in the management of the AP-Madrid application. The assessment was peer-reviewed, and discrepancies were resolved by consensus.

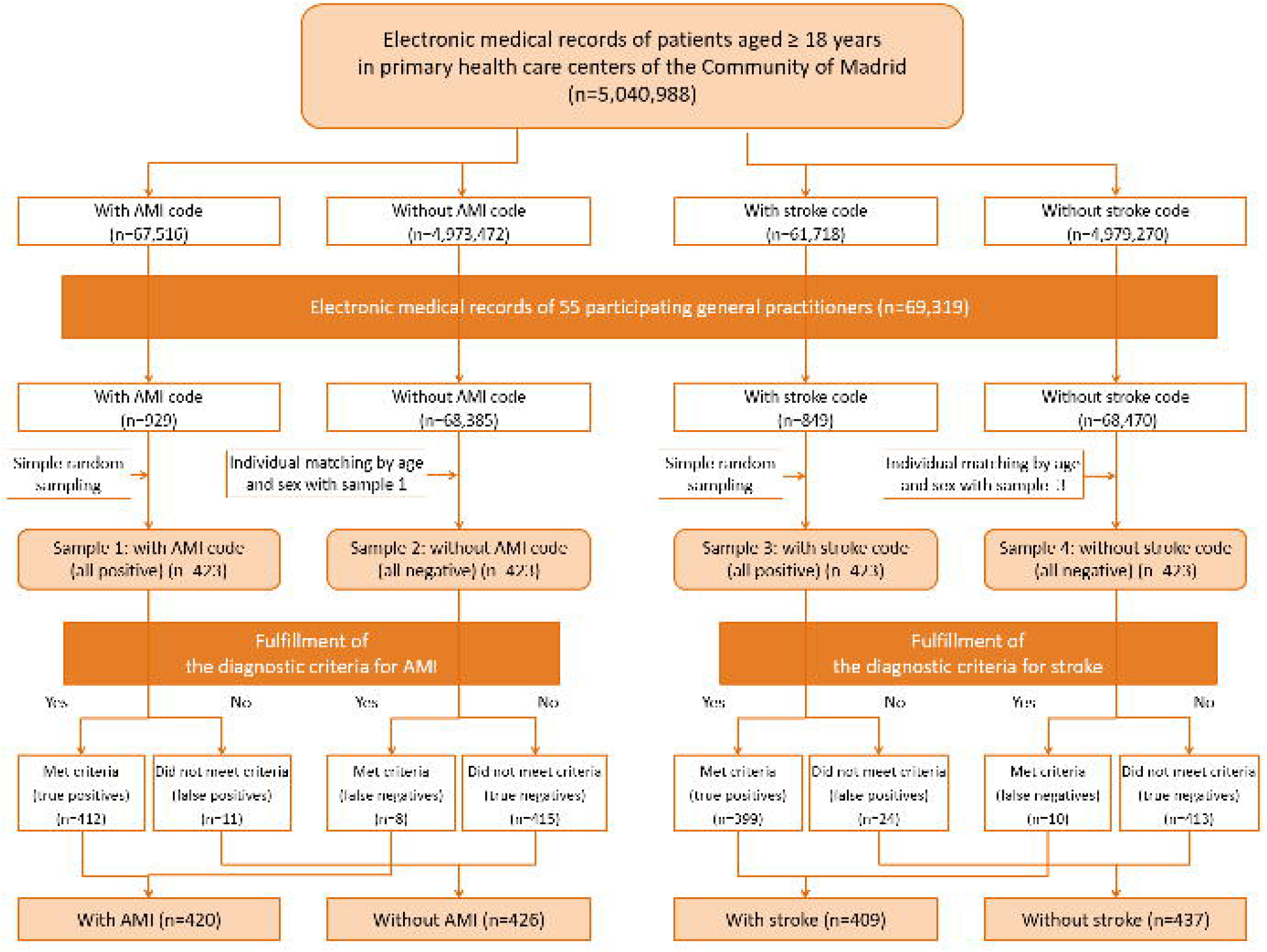

### Statistical analysis

First, a descriptive analysis of the study populations and samples was performed. The quantitative variables are expressed as the mean and standard deviation, and age is expressed as the median and interquartile range (IQR). Qualitative variables were summarized with their relative frequency.

Sensitivity and specificity, positive predictive value (PPV), and negative predictive value (NPV) with their 95% confidence intervals (CI) were calculated overall and stratified by sex and age groups. We tested whether the sensitivity and specificity differed according to the different categories of the variables using the χ^2^ test for homogeneity of the contrast statistic. When the conditions for its application were not met (any expected frequency less than 5), a two-sided Fisher’s exact test was used.

Sensitivity is the proportion of cases with AMI or stroke codes in the EMR among all cases where the diagnostic criteria could be verified. Specificity is the proportion of cases with no AMI or stroke code among those who did not meet the diagnostic criteria.

The proportion of individuals with a disease code in the EMR (apparent prevalence) should not be used as an estimate of the prevalence of a disease in that population, given that the sensitivity and specificity of these diagnoses are usually less than 100%. Thus, the proportion of individuals with a positive result includes false positives and excludes false negatives. Consequently, estimating the true prevalence of a disease requires an adjustment for misclassification resulting from the sensitivity and specificity. In this study, the formula proposed by Rogan and Gladen (13) was used for this adjustment, as follows: true prevalence = (apparent prevalence+specificity-1)/(sensitivity+specificity-1).

The degree of overall agreement between the recorded diagnosis and the reference standard, as well as the interobserver agreement, was determined using the kappa index and its CIs. According to this value, agreement is considered poor (≤ 0.20), low (0.21-0.40), moderate (0.41-0.60), good (0.61-0.80), or very good (≥ 0.81) (14).

The statistical analysis was performed using SPSS 19.0®, and the CIs of the kappa index and the predictive values were calculated with the macros for SPSS of the Laboratory of Applied Statistics of the Autonomous University of Barcelona (Spain), !KAPPA and !DT, respectively (15) (16).

### Ethics statement

To guarantee the confidentiality of the data in the validation process, we acted under the provisions of the Spanish “Organic Law on Personal Data Protection and guarantee of digital rights” and the provisions of Regulation (EU) 2016/679 of the European Parliament and of the Council of 27 April 2016 on Data Protection.

The e-MADVEVA Study protocol was approved by the Ethics Committee of Hospital Universitario Ramon y Cajal, Madrid (code 345) and the Central Research Commission of the Madrid Primary Care Management (code 02/2015). The Ethics Committee did not require informed consent because the research was performed with secondary data. In addition, all evaluators signed a confidentiality clause.

### Patient and Public Involvement

The data used are secondary without interaction with the patients. Therefore, it was neither appropriate nor possible to involve patients or the public in the design, conduct, reporting and dissemination plans of our research.

## Results

The main demographic characteristics of the population of patients over 18 years of age seen in primary care centres of the Community of Madrid with episodes of AMI (ICPC-2 K75) and stroke (ICPC-2 K90) in their EMR, as well as those of the samples selected, are described in Table 1.

**Table 1.** Main demographic characteristics of the patients aged ≥ 18 years attended in PHC centers and of samples selected.

| | N(%) | Age (years)median<br>(IQR) | Age $\geq 70$<br>years(%) | Female<br>sex(%) |
| --- | --- | --- | --- | --- |
| **Patients with diagnostic code for AMI (K-75)** | 67,516 (1.34) | 71.04 (61.31-80.54) | 52.78 | 23.20 |
| **Sample of patients with diagnostic code for AMI (sample 1)** | 423 (0.63) | 73.07 (64.02-83.12) | 59.34 | 28.37 |
| – Correct diagnosis (TP) | 412 (97.40) | 72.81 (63.93 - 82.99) | 58.74 | 27.43 |
| – Incorrect diagnosis (FP) | 11 (2.60) | 85.25 (75.85 - 90.14) | 81.82 | 63.64 |
| **Patients without diagnostic code for AMI (K-75)** | 4,973,472 (98.66) | 47.40 (36.60-61.48) | 14.81 | 53.04 |
| **Sample of patients without diagnostic code for AMI (sample 2)** | 423 (0.01) | 73.28 (63.88-82.80) | 59.10 | 28.37 |
| – Correct diagnosis (TN) | 415 (98.11) | 73.08 (63.76-82.64) | 58.80 | 28.67 |
| – Incorrect diagnosis (FN) | 8 (1.89) | 84.70 (69.89 - 86.13) | 75.00 | 12.50 |
| **Patients with diagnostic code for stroke (K-90)** | 61,718 (1.22) | 74.99 (63.99-83.81) | 62.74 | 47.19 |
| **Sample of patients with diagnostic code for stroke (sample 3)** | 423 (0.69) | 75.24 (64.00 - 83.83) | 64.07 | 46.57 |
| – Correct diagnosis (TP) | 399 (94.33) | 75.66 (64.00 - 83.83) | 64.66 | 45.86 |
| – Incorrect diagnosis (FP) | 24 (5.67) | 72.68 (62.12 - 83.99) | 54.17 | 58.33 |
| **Patients without diagnostic code for stroke (K-90)** | 4,979,270 (98.78) | 47.44 (36.62-61.48) | 14.73 | 52.71 |
| **Sample of patients with diagnostic code for stroke (sample 4)** | 423 (0.01) | 75.30 (63.88 - 84.07) | 63.83 | 46.57 |
| – Correct diagnosis (TN) | 413 (97.64) | 75.27 (63.76 - 84.02) | 63.20 | 46.73 |
| – Incorrect diagnosis (FN) | 10 (2.36) | 78.84 (71.98 - 85.16) | 90.00 | 40.00 |

AMI: acute myocardial infarction; FN: false negative; FP: false positive; IQR: interquartile range; PHC: primary health care; TN: true negative; TP: true positive

Of the 5,040,988 patients aged ≥18 years with an active medical history and regular users of primary care centres, 47.36% were male, with a mean age of 49.82 (SD 17.74).

An AMI code was identified in the EMR in 1.34% of the population; of these, 76.8% were male, with a mean age of 70.45 (SD 12.84). On the other hand, patients with an AMI code in sample 1 had a mean age of 72.70 (SD 12.3) years, and 71.63% were male.

In the same population, a diagnosis of stroke was found in the EMR in 1.22%; of these, 52.81% were male, with a mean age of 72.72 (SD 14.56) years. A total of 53.43% of the patients included in sample 3 (with a stroke code) were male, with a mean age of 73.62 (SD 13.25).

As shown in Table 2, the diagnosis of AMI was confirmed in 98.11% of cases (sensitivity), and no significant differences were found after stratifying by age group and sex. However, 97.42% of those with no recorded diagnosis of AMI did not meet the criteria (specificity); this value was 4.23% higher among males than among females.

**Table 2.** Predictive values, sensitivity, specificity, and agreement for acute myocardial infarction and stroke.

| | TP | FP | FN | TN | PPV<br>(95% CI) | NPV<br>(95% CI) | Sensitivity %<br>(95% CI) | $\chi^2$<br>p-value | Specificity %<br>(95% CI) | $\chi^2$<br>p-value | Inter-observer<br>agreement<br>kappa (95% CI) | Diagnostic<br>agreement<br>kappa (95% CI) |
| --- | --- | --- | --- | --- | --- | --- | --- | --- | --- | --- | --- | --- |
| **AMI** | 412 | 11 | 8 | 415 | 97.40<br>(95.40 – 98.54) | 98.11<br>(96.31 – 99.04) | 98.11<br>(96.29 – 99.03) |  | 97.42<br>(95.44 – 98.55) |  | 0.838<br>(0.711 – 0.966) | 0.955<br>(0.935 – 0.975) |
| – Female | 113 | 7 | 1 | 119 | 97.69<br>(95.31 - 98.88) | 99.17<br>(95.43 – 99.85) | 99.12<br>(95.20 – 99.85) | 0.689* | 94.44<br>(88.98 – 97.28) | 0.019* | 0.931<br>(0.797 – 1) | 0.933<br>(0.888 – 0.979) |
| – Male | 299 | 4 | 7 | 296 | 98.68<br>(96.66 – 99.49) | 97.69<br>(95.31 - 98.88) | 97.71<br>(95.35 - 98.89) |  | 98.67<br>(96.62 - 99.48) |  | 0.778<br>(0.590 - 0.967) | 0.964<br>(0.942 - 0.985) |
| – Age <70 | 170 | 2 | 2 | 171 | 98.84<br>(95.86 – 99.68) | 98.84<br>(95.88 – 99.68) | 98.84<br>(95.86 – 99.68) | 0.480* | 98.84<br>(95.88 – 99.68) | 0.212* | 0.887<br>(0.669 – 1) | 0.977<br>(0.954 – 0.999) |
| – Age ≥70 | 242 | 9 | 6 | 244 | 95.62<br>(92.32 - 97.54) | 97.6<br>(94.86 - 98.90) | 97.58<br>(94.82 - 98.89) |  | 96.44<br>(93.38 - 98.12) |  | 0.823<br>(0.670 - 0.975) | 0.940<br>(0.910 - 0.970) |
| **Stroke** | 399 | 24 | 10 | 413 | 94.33<br>(91.70 - 96.16) | 97.64<br>(95.70 - 98.71) | 97.56<br>(95.56 - 98.68) |  | 94.51<br>(91.96 - 96.28) |  | 0.862<br>(0.778 - 0.946) | 0.920<br>(0.893 - 0.946) |
| – Female | 183 | 14 | 4 | 193 | 92.89<br>(88.43 - 95.72) | 97.97<br>(94.90 - 99.21) | 97.86<br>(94.63 – 99.17) | 0.760 * | 93.24<br>(88.97 – 95.93) | 0.268 | 0.915<br>(0.819 – 1) | 0.909<br>(0.867 – 0.950) |
| – Male | 216 | 10 | 6 | 220 | 95.58<br>(92.05 – 97.58) | 97.35<br>(94.33 – 98.78) | 97.30<br>(94.23 – 98.76) |  | 95.65<br>(92.18 – 97.62) |  | 0.813<br>(0.677 – 0.948) | 0.929<br>(0.895 – 0.963) |
| – Age <70 | 141 | 11 | 1 | 152 | 92.76<br>(87.51 – 95.91) | 99.35<br>(96.39 – 99.89) | 99.30<br>(96.12 – 99.88) | 0.175 * | 93.25<br>(88.32 - 96.19) | 0.374 | 0.913<br>(0.794 - 1) | 0.921<br>(0.878 - 0.965) |
| – Age ≥70 | 258 | 13 | 9 | 261 | 95.36<br>(92.22 - 97.27) | 96.67<br>(93.79 - 98.24) | 96.63<br>(93.92 - 98.28) |  | 95.26<br>(92.05 - 97.21) |  | 0.838<br>(0.73 - 0.95) | 0.919<br>(0.887 - 0.953) |

AMI: acute myocardial infarction; 95% CI: 95% confidence interval; FN: false negative; FP: false positive; NPV: negative predictive value; PPV: positive predictive value; TN: true negative; TP: true positive; Two-sided Fisher's exact test.

In seven of the 11 cases categorized as false positives where the diagnosis could not be confirmed, this was unstable angina with severe 2- or 3-vessel disease; in four of the cases, the original label had been changed from AMI to unstable angina. In three EMRs, the physician also reported that the patient had an AMI, but the information could not be verified. Furthermore, atypical pain was reported in another case, in which a cardiologist ruled out the diagnosis.

In seven of the eight false negatives, the description of angina had been altered, either by changing it or adding the AMI label, as five cases had previously had episodes of angina. Reference to an AMI in a cardiology report in 1992 was found in another EMR.

The sensitivity of a diagnosis of stroke was 97.56%, and the specificity was 94.51%, with no statistically significant differences found for the different sex and age strata.

In 17 of the 24 false positives, GPs changed the label without changing the code (nine transient ischemic attacks, five seizures, one meningioma, one hypertensive crisis, and one vasectomy). In five other cases with labels where the diagnosis could not be confirmed, these were transient ischemic attacks. In two EMRs, no data were found to cross-check the diagnosis.

The label had been changed in nine of the ten false negatives; in seven cases, the change was from transient ischemic attacks to stroke, replacing or adding to the existing label. In another EMR, an emergency department report referring to a stroke was found.

The overall agreement between the diagnosis recorded in the EMR and the reference standard, measured as the kappa concordance index, was very good for diagnosing AMI (κ = 0.955) and stroke (κ = 0.920).

The overall degree of agreement between observers, as measured by the kappa index, is very good for the overall diagnoses of AMI (κ = 0.838) and stroke (κ = 0.862) and for the different strata of the variables sex and age over 69 years. In all cases, κ indices above 0.880 were achieved.

The true prevalence of AMI in the population aged ≥18 years in the Community of Madrid was 1.38% (0.57% in women and 2.24% in men). This increased progressively with age, reaching 5.5% in those over 80 (3.23% in women and 9.77% in men).

The true prevalence of stroke was 1.27% (1.13% in women and 1.42% in men); in the group aged over 80 years, the prevalence of stroke was 7.35% (3.23% in women and 9.77% in men).

Table 3 shows the differences in the prevalence of AMI and stroke, as recorded in the EMR (apparent prevalence) and according to the reference standard (true prevalence), both overall and stratified by age group and sex.

**Table 3.** Apparent prevalence (diagnosis in the EMR) and true prevalence (fulfillment of the diagnostic criteria) of AMI and stroke in persons aged ≥ 18 years in primary health care centers of the Community of Madrid.

| ALL |  |  |  |  | FEMALE |  |  |  | MALE |  |  |  |
| --- | --- | --- | --- | --- | --- | --- | --- | --- | --- | --- | --- | --- |
| AMI | N | n | Apparent prevalence (95% CI) | True prevalence (95% CI) | N | n | Apparent prevalence (95% CI) | True prevalence (95% CI) | N | n | Apparent prevalence (95% CI) | True prevalence (95% CI) |
| **Total** | 5,040,988 | 67,516 | 1.34<br>(1.33 - 1.35) | 1.38<br>(1.37 - 1.39) | 2,653,677 | 15,663 | 0.59<br>(0.58 - 0.6) | 0.57 (0.56 - 0.58) | 2,387,311 | 51,853 | 2.17<br>(2.15 - 2.23) | 2.24<br>(2.22 - 2.26) |
| 18-50 y | 2,761,166 | 4,027 | 0.15<br>(0.14 - 0.15) | 0.14<br>(0.13 - 0.14) | 1,395,907 | 801 | 0.06<br>(0.05 - 0.06) | 0<br>(0 - 0) | 1,365,259 | 3,226 | 0.24<br>(0.23 - 0.24) | 0.23<br>(0.22 - 0.24) |
| 51-60 y | 880,580 | 10,945 | 1.24<br>(1.22 - 1.27) | 1.28<br>(1.25 - 1.3) | 456,252 | 1,656 | 0.36<br>(0.35 - 0.38) | 0.33<br>(0.31 - 0.35) | 424,328 | 9,289 | 2.19<br>(2.15 - 2.23) | 2.26<br>(2.21 - 2.3) |
| 61-70 y | 626,926 | 16,908 | 2.7<br>(2.66 - 27.37) | 2.78<br>(2.74 - 2.83) | 338,843 | 2,706 | 0.8<br>(0.77 - 0.83) | 0.79<br>(0.76 - 0.82) | 288,083 | 14,202 | 4.93<br>(4.85 - 5.01) | 5.1<br>(5.02 - 5.18) |
| 71-80 y | 436,045 | 17,766 | 4.07<br>(4.02 - 4.13) | 4.21<br>(4.15 - 4.27) | 244,789 | 3,790 | 1.55<br>(1.5 - 1.6) | 1.6<br>(1.55 - 1.65) | 191,256 | 13,976 | 7.31<br>(7.19 - 7.43) | 7.57<br>(7.45 - 7.69) |
| >80 y | 336,271 | 17,870 | 5.31<br>(5.24 - 5.39) | 5.5<br>(5.42 - 5.58) | 217,886 | 6,710 | 3.08<br>(3.01 - 3.15) | 3.23<br>(3.16 - 3.31) | 118,385 | 11,160 | 9.43<br>(9.26 - 9.59) | 9.77<br>(9.6 - 9.94) |
| Stroke | N | n | Apparent prevalence (95% CI) | True prevalence (95% CI) | N | n | Apparent prevalence (95% CI) | True prevalence (95% CI) | N | n | Apparent prevalence (95% CI) | True prevalence (95% CI) |
| **Total** | 5,040,988 | 61,718 | 1.22<br>(1.21 - 1.23) | 1.27<br>(1.26 - 1.28) | 2,653,677 | 29,123 | 1.1<br>(1.09 - 1.11) | 1.13<br>(1.12 - 1.14) | 2,387,311 | 32,595 | 1.37<br>(1.35 - 1.38) | 1.42<br>(1.41 - 1.44) |
| 18-50 y | 2,761,166 | 4,894 | 0.18<br>(0.17 - 0.18) | 0.13<br>(0.13 - 0.14) | 1,395,907 | 2,239 | 0.16<br>(0.15 - 0.17) | 0.1<br>(0.1 - 0.11) | 1,365,259 | 2,655 | 0.19<br>(0.19 - 0.2) | 0.16<br>(0.16 - 0.17) |
| 51-60 y | 880,580 | 6,764 | 0.77<br>(0.75 - 0.79) | 0.77<br>(0.76 - 0.79) | 456,252 | 2,534 | 0.56<br>(0.53 - 0.58) | 0.54<br>(0.51 - 0.56) | 424,328 | 4,230 | 1<br>(0.97 - 1.03) | 1.03<br>(1 - 1.06) |
| 61-70 y | 626,926 | 11,339 | 1.81<br>(1.78 - 1.84) | 1.9<br>(1.87 - 1.94) | 338,843 | 4,296 | 1.27<br>(1.23 - 1.31) | 1.32<br>(1.28 - 1.36) | 288,083 | 7,043 | 2.44<br>(2.39 - 2.5) | 2.58<br>(2.53 - 2.64) |
| 71-80 y | 436,045 | 15,784 | 3.62<br>(3.56 - 3.68) | 3.87<br>(3.82 - 3.93) | 244,789 | 6,718 | 2.74<br>(2.68 - 2.81) | 2.94<br>(2.87 - 3.01) | 191,256 | 9,066 | 4.74<br>(4.65 - 4.84) | 5.05<br>(4.96 - 5.15) |
| >80 y | 336,271 | 22,937 | 6.82<br>(6.74 - 6.91) | 7.35<br>(7.26 - 7.44) | 217,886 | 13,336 | 6.12<br>(6.02 - 6.22) | 6.64<br>(6.54 - 6.75) | 118,385 | 9,601 | 8.11<br>(7.96 - 8.27) | 8.68<br>(8.52 - 8.84) |

AMI: acute myocardial infarction; Apparent prevalence: proportion of individuals with a disease code in the EMR; EMR: electronic medical record; True prevalence: (apparent prevalence+specificity-1)/(sensitivity+specificity-1); y: years.

## Discussion

The results of the e-MADVEVA Study showed very good agreement with the reference standard and high sensitivity and specificity overall and in each sex category and age group.

These results are consistent with those of other published studies, although most validate the coding of these diagnoses in hospital registries or specific registries using ICD-9 or ICD-10 codes.

Thus, for the diagnosis of AMI, the systematic review by McCormick et al. (17) of 30 studies published between 1984 and 2010 found that sensitivity was, in most cases, higher than 86%, specificity higher than 89%, PPV higher than 93%, and NPV higher than 75%. In their review of 31 studies published between 2000 and 2014, Rubbo et al (18) found PPVs greater than 70%.

In the case of stroke, our results are superior to those found in other studies, such as that of Baldereschi et al. (19) in Italy (sensitivity 70.54%, PPV 97.3%), Hall et al. (20) in Canada (sensitivity 82.2%, PPV 68.8%), Johnsen et al. (21) in Denmark (PPV 87.6%), and Porter et al. in Canada (22) (sensitivity 97.3%). In a systematic review of 77 studies published between 1976 and 2015 by McCormick et al. (23), the sensitivity of ICD-9 and ICD-10 codes for any cerebrovascular disease was over 82% in most studies, and the PPV was over 81%.

The specificity found in our study was 94.51%, and the NPV was 97.64%, similar to those found by Baldereschi et al. (19) (95.56% and 85.64%, respectively) and by McCormick et al. (23) (both above 95%).

Most epidemiological studies on AMI and stroke in Spain estimate incidence, hospital admissions, and mortality (24) (25) (26) (27) (28). The few prevalence studies found show great variability with respect to terminology, definition, and methodology, as well as in the age groups assessed (29) (30) (31).

The adult questionnaire of the 2017 National Health Survey of the Spanish National Institute of Statistics includes the question “Has a doctor ever diagnosed you with an acute myocardial infarction?” and “Has a doctor ever diagnosed you with stroke (embolism, cerebral infarction, cerebral hemorrhage)?” (32). Figure 2 compares the prevalence of AMI and stroke found in this study with the rates of positive responses to these questions by sex and age group. As can be seen, the results are very similar.

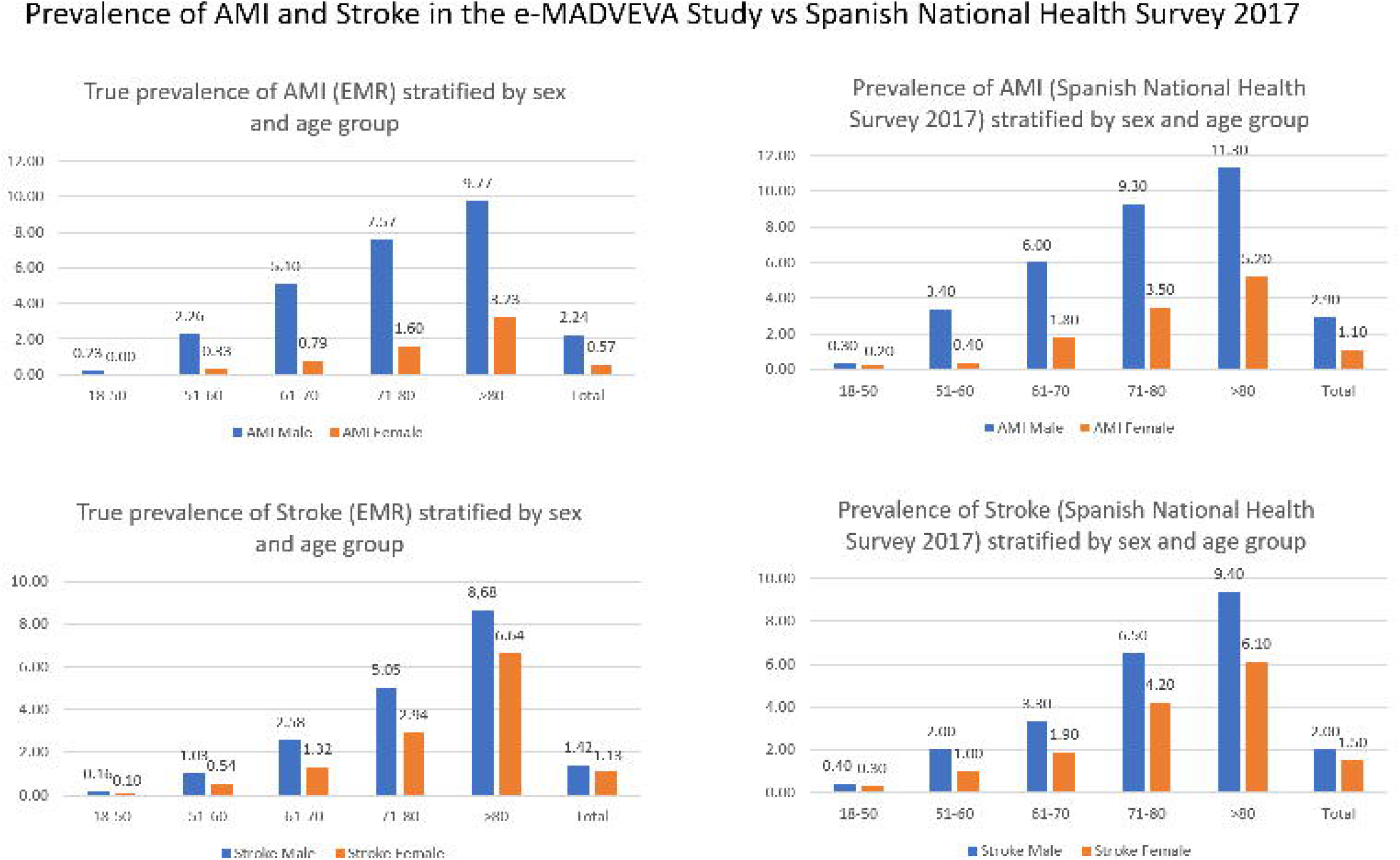

It is noteworthy that most errors in the diagnosis of AMI (both false positives and false negatives) are due to coding errors for unstable angina episodes and, in the case of stroke, for TIA episodes, which could be considered earlier stages of both diseases. In addition, changes made by clinicians in episode labeling have been detected primarily in older diagnoses, suggesting that GPs tend to make fewer coding errors over time.

The major strength of the e-MADVEVA Study is the individual validation (manual validation) of EMR by comparing each case and non-case, matched by age, with an accepted reference gold standard. The validation method allows for calculating PPV, NPV, Sensitivity, and Specificity, in contrast with other methods such as questionnaires for healthcare practitioners or patients and comparing rates in a comparable population (33).

Our study is limited by the fact that the study design makes it impossible for researchers to remain blind to the medical records. Blinding could have led to more remarkable agreement than would have been the case if they had been blind to this information. In addition, in-hospital mortality due to AMI and stroke may not have been recorded in primary care EMRs, with the result that the prevalence of patients who die in hospital due to a first AMI or stroke may be underestimated.

The gold standard of our study was the fulfillment of diagnostic criteria that can be verified with information collected in the primary care EMR. For this reason, we cannot assume that false positives are diagnostic errors, only that they could not be verified, thus potentially leading us to underestimate prevalence.

We selected episodes using codes, regardless of the labels. This entails a potential selection bias in cases where the professional recording the episode modifies the label. However, only 0.26% and 0.74% of the AMI and stroke codes, respectively, had a label that did not correspond to the episode. Conversely, 0.07% and 0.11% of the labels of patients without cardiovascular episode codes were modified and corresponded to AMI and stroke, respectively.

Lastly, the ICPC-2 codes studied for AMI (ICPC-2 K75) and stroke (ICPC-2 K90) do not allow differentiation between the different types of AMI and stroke and, therefore, cannot be used in studies requiring discrimination between types.

## Conclusions

The validation results show that diagnoses of AMI and stroke recorded in primary care EMRs are a helpful tool for epidemiological studies.

The prevalence of AMI and stroke was lower than 2% in the population aged over 18 years. However, the prevalence of both diseases increases progressively with age.

## Data Availability

The datasets generated during the e-MADVEVA Study are available from the corresponding author on reasonable request.

## Licence statement

I, the Submitting Author has the right to grant and does grant on behalf of all authors of the Work (as defined in the below author licence), an exclusive licence and/or a non-exclusive licence for contributions from authors who are: i) UK Crown employees; ii) where BMJ has agreed a CC-BY licence shall apply, and/or iii) in accordance with the terms applicable for US Federal Government officers or employees acting as part of their official duties; on a worldwide, perpetual, irrevocable, royalty-free basis to BMJ Publishing Group Ltd (“BMJ”) its licensees and where the relevant Journal is co-owned by BMJ to the co-owners of the Journal, to publish the Work in BMJ Open and any other BMJ products and to exploit all rights, as set out in our licence.

The Submitting Author accepts and understands that any supply made under these terms is made by BMJ to the Submitting Author unless you are acting as an employee on behalf of your employer or a postgraduate student of an affiliated institution which is paying any applicable article publishing charge (“APC”) for Open Access articles. Where the Submitting Author wishes to make the Work available on an Open Access basis (and intends to pay the relevant APC), the terms of reuse of such Open Access shall be governed by a Creative Commons licence – details of these licences and which Creative Commons licence will apply to this Work are set out in our licence referred to above.

## Consent for publication

Not applicable

## Authors’ contributions

CBL conceived the study, and MASF, ICG, MSP, and CBL developed its design. CBL, MASF, JAH, and JCV contributed to data acquisition. JCV, VIC, and PGC coordinated the research group. CBL, MSP, VIC, and ALA performed the statistical analysis. All authors contributed to the interpretation of results. CBL worked with MASF to develop the first draft of the manuscript. All authors contributed to revisions of the manuscript and the final content. All authors have approved the final manuscript, take responsibility for parts of the content, and have agreed to be accountable for all aspects of the work.

## Acknowledgments

We thank Thomas O’Boyle for writing assistance.

## Abbreviations

AMI: Acute myocardial infarction
EMR: Electronic medical records
ESC/ACCF/AHA/WHF: European Society of Cardiology, the American College of Cardiology Foundation, American Heart Association and the World Heart Federation
ICD-9: International Classification of Diseases, Ninth Revision
ICD-10: International Classification of Diseases, Tenth Revision
ICPC-2: Second edition of the International Classification of Primary Care
CI: Confidence interval
PPV: Positive predictive value
NPV: Negative predictive value

